# Association of non-HDL-C and risk of incident hypertension

**DOI:** 10.1101/2025.01.14.25320570

**Authors:** Xinxin He, Lei Yin, Yujie Zhao, Bin Yang, Yang Zhao, Xueru Fu, Yuying Wu, Mengdi Wang, Mengna Liu, Yijia Su, Yongcheng Ren, Yu Liu, Ming Zhang, Dongsheng Hu

## Abstract

**Background:** The study aimed to investigate the association between non-high-density lipoprotein cholesterol (non-HDL-C) and risk of incident hypertension, especially concerning the association of different levels of low-density lipoprotein cholesterol (LDL-C) and non-HDL-C with risk of incident hypertension.

**Methods:** A total of 10,623 participants from the Rural Chinese Cohort Study were included in the analyses. Non-HDL-C is the total cholesterol in the blood minus high-density lipoprotein cholesterol (HDL-C). The odds ratios (ORs) and 95% confidence intervals (95% CIs) between non-HDL-C, LDL-C, and risk of incident hypertension were estimated using logistic regression models.

**Results:** During the 10-year follow-up, 3,110 newly diagnosed hypertensive patients were identified. After adjustment for potential confounders, baseline non-HDL-C level was associated with higher risk of hypertension: the OR for the highest vs lowest quartile was 1.79 (95% CI: 1.43-2.24). For each standard deviation (SD) increase in non-HDL-C, risk of incident hypertension increased by 41%. When LDL-C was within the ideal level (<2.6 mmol/L) but non-HDL-C was above the ideal level (≥3.4 mmol/L), the risk of incident hypertension was elevated by 56% compared to when both of them were at ideal levels. Conversely, no such increased risk was observed when the LDL-C level was non-ideal while non-HDL-C remained within the ideal level.

**Conclusions:** Serum non-HDL-C level was found to be positively associated with the risk of incident hypertension in rural Chinese adults, an association independent of LDL-C, suggesting that further lowering of non-HDL-C by interventions to reduce residual risk of incident hypertension would be beneficial.

## Introduction

Due to population aging, lifestyle changes, and longer life expectancies, the worldwide prevalence of hypertension has been on a consistent rise over the last 20 to 30 years. The count of individuals aged between 30 and 79 suffering from hypertension globally increased from 648 million to 1.28 billion over the period from 1990 to 2019 ^1^. About 256.7 million adults in China between the ages of 30 and 79 had hypertension in 2019, with an overall age-standardized prevalence of 27% ^2^. Alongside this, a high blood pressure level increases the risk of a number of diseases, collectively causing nearly 8.5 million deaths each year ^3,4^. Identifying risk factors for risk of incident hypertension, early detection, and managing blood pressure are essential due to the condition’s widespread occurrence and heavy impact on health.

A significant contributor to hypertension is dyslipidemia, a condition marked by elevated concentrations of total cholesterol (TC), low-density lipoprotein cholesterol (LDL-C), and non-high-density lipoprotein cholesterol (non-HDL-C) ^5,6^. Although statins are predominantly prescribed to decrease LDL-C concentrations, the possibility of cardiovascular diseases (CVDs) persists at heightened levels even when LDL-C is kept within the prescribed targets ^7^. Non-HDL-C might be another indicator within the lipid profile that contributes to the development of CVDs, according to recent studies ^8,9^. Non-HDL-C is the total cholesterol (TC) in the blood minus high-density lipoprotein cholesterol (HDL-C) ^10,11^. Launched by the American Heart Association (AHA) in 2022, Life’s Essential 8 modernizes the idea of cardiovascular health, particularly with regard to lipid testing, suggesting non-HDL-C as a crucial indicator ^12^.

Hypertension is a primary risk factor for nearly all types of CVDs that individuals may develop during their lifetime ^13^. Previous studies have shown that higher non-HDL-C levels increase the risk of incident hypertension for individuals compared to those with lower levels ^5,14^. The aim of this study, therefore, was to investigate the relationship of non-HDL-C with risk of incident hypertension while further assessing the relationship of varying LDL-C and non-HDL-C levels with risk of incident hypertension in an effort to furnish further epidemiological evidence for the proactive prevention of this condition.

## Materials and Methods

### Study population

The Rural Chinese Cohort Study is a prospective cohort study that utilized cluster random sampling to recruit participants from a rural area within Henan Province, China ^15^. Participants were at least 18 years old and free of serious mental illnesses, physical impairments, dementia, Alzheimer’s disease, tuberculosis, HIV/AIDS, or other infectious diseases ^15^. The study’s timeline included a baseline survey from July-August 2007 to July-August 2008, a first follow-up survey from July-August 2013 to July-October 2014 with an 85.5% follow-up rate, and a second follow-up survey from July-August 2018 to July-August 2020 with a 92.8% follow-up rate ^16^. Details about the study’s participants, design, and data collection have already been provided ^15^.

20,194 individuals were enrolled in total at baseline. Participants in this study were not included if they were: (1) people with baseline hypertension (n = 6,299); (2) taking lipid-lowering drugs at baseline (n = 433); (3) absent of baseline TC, HDL-C, and LDL-C data (n = 344); or (4) their hypertension status was unknown during the 10-year follow-up (n = 2,495) (Supplementary Figure 1).

### Baseline data collection

A team of trained investigators used laboratory measurements, anthropometry, physical examinations, and in-person questionnaire interviews to collect pertinent data. The questionnaire contained a range of socio-demographic attributes, including age, gender, occupation, education, marital status, and lifestyles. Additionally, it covered personal medical histories pertaining to blood pressure, blood glucose, and lipid-lowering pharmaceuticals, and the family history of diseases ^15^. Smoking was defined as consuming 100 or more cigarettes during one’s lifetime ^17^. Drinking was defined as consuming alcohol more than 12 times in the previous 12 months ^18^. The diagnostic criteria for dyslipidemia encompass the following: TC ≥5.2 mmol/L, LDL-C ≥3.4 mmol/L, triglycerides (TG)≥1.7 mmol/L, HDL-C<1.00 mmol/L, and non-HDL-C ≥4.1 mmol/L. Meeting one or more of these criteria is sufficient for a diagnosis of dyslipidemia ^19^. The International Physical Activity Questionnaire was used to classify physical activity into three levels: low, medium, and high ^20^. The heights of the participants were recorded with precision to the nearest 0.1 cm as they stood upright. Their weights were similarly assessed to the closest 0.5 kg using a vertical weight scale ^21^.

After a minimum fast of 8 hours, blood specimens were collected, and the Hitachi 7060 Automatic Analyzer was used to assess fasting plasma glucose (FPG), TG, TC, HDL-C, and LDL-C. The non-HDL-C level was computed using the formula:

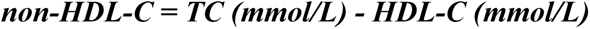

### Classification criteria for LDL-C and non-HDL-C

We categorized LDL-C and non-HDL-C using uniform optimal concentration thresholds, in line with the Chinese Guideline for Lipid Management (Primary Care Version 2024), as well as the AHA’s “Life’s Essential 8” guidelines ^12,19^. It is ideal to have LDL-C level<2.6 mmol/L and non-HDL-C level<3.4 mmol/L. Accordingly, we categorized the study population into 4 different groups: those with LDL-C<2.6 mmol/L and non-HDL-C<3.4 mmol/L as the control group, those with LDL-C<2.6 mmol/L and non-HDL-C ≥3.4 mmol/L, those with LDL-C ≥2.6 mmol/L and non-HDL-C<3.4 mmol/L, and those with both LDL-C ≥2.6 mmol/L and non-HDL-C ≥3.4 mmol/L. The study aim was to assess the risk of incident hypertension within each group relative to the control, with a particular emphasis on the instances where LDL-C or non-HDL-C were not optimal.

### Outcomes assessment

Thirty minutes before the blood pressure measurement, the participants had to empty their bladders, avoid caffeine, alcohol, and smoking, and sit still for five minutes before the measurement, with their elbows at heart level during the measurement. A trained health worker utilized an electronic blood pressure monitoring device (HEM-770 AFuzzy) to conduct three measurements spaced 30 seconds apart, in line with the American Heart Association’s recommended protocol ^22^. If any two of the three measurements differed by more than 5 mmHg, they were re-measured, with the average of the measured values then used for analysis. Hypertension is defined by a systolic blood pressure of ≥140 mmHg, a diastolic blood pressure of ≥90 mmHg, or use of antihypertensive medication ^23^.

Follow-up measures employed the same strategies as at baseline, using the same definition of hypertension.

### Statistical analysis

The population was divided into four groups (Q1 to Q4) based on lower quartile, median, and upper quartile of non-HDL-C levels at baseline, representing the lowest (Q1), lower middle (Q2), upper middle (Q3), and highest (Q4) levels, and participants in each quartile were compared with participants in the lowest quartile. In this study, all continuous variables exhibited skewness, thus their distributional properties were characterized by the median (interquartile range from the upper to the lower quartile). To assess group differences, the Wilcoxon rank-sum test and the Kruskal-Wallis test were employed. Categorical variables were summarized by frequency counts and percentages, with group comparisons conducted using the *χ*^2^ test. The odds ratios (ORs) and 95% confidence intervals (95% CIs) for non-HDL-C and risk of incident hypertension, and for various levels of LDL-C and non-HDL-C and risk of incident hypertension, were calculated using logistic regression models. Further, 4 models that accounted for baseline confounders were established. Model 1 did not adjust; model 2 adjusted for age and gender; and model 3 adjusted for marital status, education, smoking, drinking, physical activity, family history of hypertension, BMI, and FPG on the basis of model 2. Further, we adjusted LDL-C based on model 3 to create model 4, with the aim of examining the independent association of non-HDL-C with risk of incident hypertension, disassociated from the influence of LDL-C. Restricted cubic splines (RCS) with 4 knots were used to explore the dose-response relationship of non-HDL-C with risk of incident hypertension.

Subgroup analyses of different subgroups (age, gender, smoking, drinking, dyslipidemia, physical activity, and family history of hypertension) were performed. In addition, sensitivity analyses were performed to further evaluate the robustness of the findings after excluding patients with baseline dyslipidemia, prehypertension, cancer, end-stage renal disease, and CVDs.

We further incorporated an interaction term for LDL-C level (<2.6 mmol/L versus ≥2.6 mmol/L) and non-HDL-C level (<3.4 mmol/L versus ≥3.4 mmol/L) into the model to assess the combined and interactive effects. The measure of multiplicative interaction was expressed as the OR and its 95% CI for the interaction term. Three metrics were employed for the additive interaction: the synergy index (SI), proportion attributable to interaction (AP), and relative excess risk caused by interaction (RERI). Specifically, RERI = 0, AP = 0, and SI = 1 imply no additive interaction between LDL-C and non-HDL-C for risk of incident hypertension. If RERI > 0, AP > 0, and SI > 1, the combined effect of LDL-C and non-HDL-C on risk of incident hypertension exceeds individual effects, suggesting synergy; however, RERI < 0, AP < 0, and SI < 1 indicate the combined effect is less than the sum of individual effects. We calculated 95% CIs for these three measures using the delta method.

Analyses were conducted using SAS 9.4 software (SAS Institute, Cary, NC, USA) and R version 4.3.0 (R Foundation). All statistical tests performed were two-tailed, and a *P*-value of less than 0.05 was deemed indicative of statistical significance.

## Results

### Baseline characteristics

During the 10-year follow-up, 3,110 newly diagnosed hypertension cases were recorded, comprising 1,247 males and 1,863 females. Age, marital status, educational level, physical activity, family history of hypertension, BMI, waist circumference (WC), SBP, DBP, TC, TG, HDL-C, LDL-C, FPG, and non-HDL-C all differed statistically significantly (*P* < 0.05) between the hypertensive and non-hypertensive groups, but gender, smoking, or drinking did not differ significantly (*P* > 0.05) (Table 1).

**Table 1.**
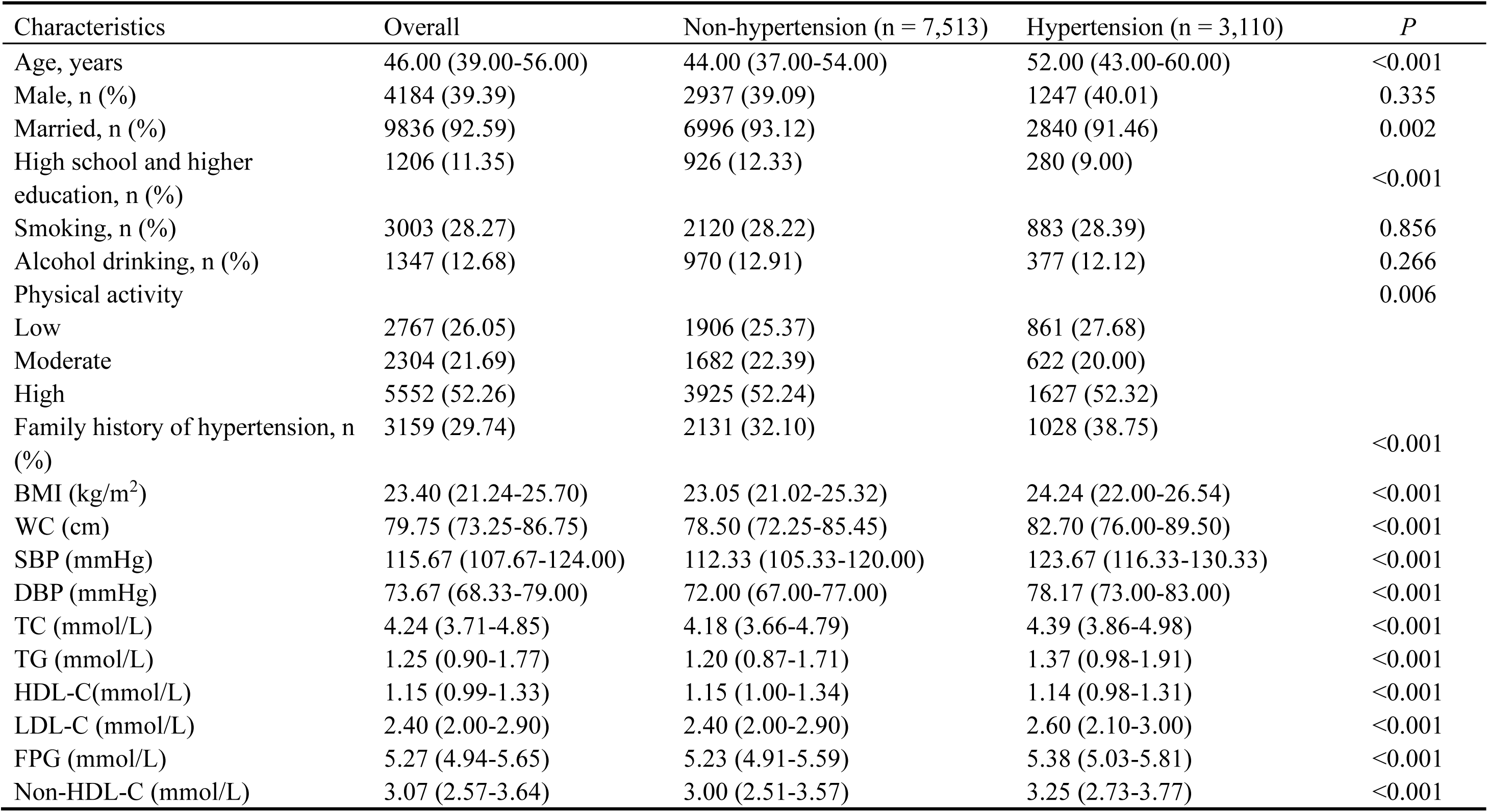
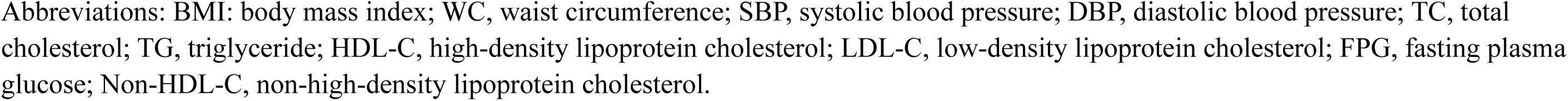
Baseline characteristics of 10,623 participants.

| Characteristics | Overall | Non-hypertension (n = 7,513) | Hypertension (n = 3,110) | *P* |
| --- | --- | --- | --- | --- |
| Age, years | 46.00 (39.00-56.00) | 44.00 (37.00-54.00) | 52.00 (43.00-60.00) | <0.001 |
| Male, n (%) | 4184 (39.39) | 2937 (39.09) | 1247 (40.01) | 0.335 |
| Married, n (%) | 9836 (92.59) | 6996 (93.12) | 2840 (91.46) | 0.002 |
| High school and higher education, n (%) | 1206 (11.35) | 926 (12.33) | 280 (9.00) | <0.001 |
| Smoking, n (%) | 3003 (28.27) | 2120 (28.22) | 883 (28.39) | 0.856 |
| Alcohol drinking, n (%) | 1347 (12.68) | 970 (12.91) | 377 (12.12) | 0.266 |
| Physical activity |  |  |  | 0.006 |
| Low | 2767 (26.05) | 1906 (25.37) | 861 (27.68) |  |
| Moderate | 2304 (21.69) | 1682 (22.39) | 622 (20.00) |  |
| High | 5552 (52.26) | 3925 (52.24) | 1627 (52.32) |  |
| Family history of hypertension, n (%) | 3159 (29.74) | 2131 (32.10) | 1028 (38.75) | <0.001 |
| BMI (kg/m<sup>2</sup>) | 23.40 (21.24-25.70) | 23.05 (21.02-25.32) | 24.24 (22.00-26.54) | <0.001 |
| WC (cm) | 79.75 (73.25-86.75) | 78.50 (72.25-85.45) | 82.70 (76.00-89.50) | <0.001 |
| SBP (mmHg) | 115.67 (107.67-124.00) | 112.33 (105.33-120.00) | 123.67 (116.33-130.33) | <0.001 |
| DBP (mmHg) | 73.67 (68.33-79.00) | 72.00 (67.00-77.00) | 78.17 (73.00-83.00) | <0.001 |
| TC (mmol/L) | 4.24 (3.71-4.85) | 4.18 (3.66-4.79) | 4.39 (3.86-4.98) | <0.001 |
| TG (mmol/L) | 1.25 (0.90-1.77) | 1.20 (0.87-1.71) | 1.37 (0.98-1.91) | <0.001 |
| HDL-C (mmol/L) | 1.15 (0.99-1.33) | 1.15 (1.00-1.34) | 1.14 (0.98-1.31) | <0.001 |
| LDL-C (mmol/L) | 2.40 (2.00-2.90) | 2.40 (2.00-2.90) | 2.60 (2.10-3.00) | <0.001 |
| FPG (mmol/L) | 5.27 (4.94-5.65) | 5.23 (4.91-5.59) | 5.38 (5.03-5.81) | <0.001 |
| Non-HDL-C (mmol/L) | 3.07 (2.57-3.64) | 3.00 (2.51-3.57) | 3.25 (2.73-3.77) | <0.001 |

Abbreviations: BMI: body mass index; WC, waist circumference; SBP, systolic blood pressure; DBP, diastolic blood pressure; TC, total cholesterol; TG, triglyceride; HDL-C, high-density lipoprotein cholesterol; LDL-C, low-density lipoprotein cholesterol; FPG, fasting plasma glucose; Non-HDL-C, non-high-density lipoprotein cholesterol.

### Association of baseline serum non-HDL-C level and risk of incident hypertension

Table 2 shows the ORs and 95% CIs for the relationship between non-HDL-C and risk of incident hypertension. The cumulative incidence of hypertension for participants in groups 1 to 4 of the non-HDL-C levels were 21.42%, 26.85%, 33.01%, and 35.68%, respectively. The OR (95% CI) for risk of incident hypertension in the top vs the bottom quartile was 2.04 (95% CI: 1.80-2.30, *P* for trend <0.001) in the unadjusted model 1. After adjustment for potential confounders, baseline non-HDL-C level was associated with higher risk of hypertension: the OR for the highest vs lowest quartile was 1.79 (95% CI: 1.43-2.24, *P* for trend <0.001, model 4). Further, the risk of incident hypertension elevated with non-HDL-C with each standard deviation (SD) increase, with an adjusted OR (95% CI) of 1.41 (1.27-1.57) in model 4. Figure 1 shows the dose-response association of non-HDL-C level and risk of incident hypertension, revealing a positive, nonlinear correlation (*P* _nonlinear_ =0.003). Compared with the non-HDL-C concentrations of <3.01 mmol/L, as the non-HDL-C increased, the adjusted OR value for risk of incident hypertension also increased radically.

**Table 2.** Association of baseline non-HDL-C level and risk of incident hypertension.

| Non-HDL-C level<br>(mmol/L) | Cases | Number of<br>participants | Cumulative<br>incidence (%) | OR (95% CI) |  |  |  |
| --- | --- | --- | --- | --- | --- | --- | --- |
|  |  |  |  | Model 1 | Model 2 | Model 3 | Model 4 |
| Q1 (<2.57) | 564 | 2633 | 21.42 | 1.00 (ref) | 1.00 (ref) | 1.00 (ref) | 1.00 (ref) |
| Q2 (2.57-3.07) | 710 | 2644 | 26.85 | 1.35 (1.19-1.53) | 1.16 (1.01-1.32) | 1.27 (1.11-1.46) | 1.33 (1.15-1.54) |
| Q3 (3.07-3.64) | 882 | 2672 | 33.01 | 1.81 (1.60-2.04) | 1.35 (1.18-1.53) | 1.45 (1.26-1.66) | 1.57 (1.33-1.86) |
| Q4 (≥3.64) | 954 | 2674 | 35.68 | 2.04 (1.80-2.30) | 1.34 (1.18-1.53) | 1.54 (1.34-1.77) | 1.79 (1.43-2.24) |
| *P* for trend |  |  |  | <0.001 | <0.001 | <0.001 | <0.001 |
| Per SD increase |  |  |  | 1.31 (1.26-1.36) | 1.13 (1.08-1.18) | 1.17 (1.11-1.22) | 1.41 (1.27-1.57) |

The population was divided into four groups (Q1 to Q4) based on lower quartile, median, and upper quartile of non-HDL-C levels at baseline.
Q1: lowest level group; Q2: lower middle level group; Q3: upper middle level group; Q4: highest level group.
Model 1: unadjusted
Model 2: adjusted for age and gender.
Model 3: model 2 + further adjusted for marital status, education, smoking, drinking, physical activity, family history of hypertension, BMI, and FPG.
Model 4: model 2 + further adjusted for marital status, education, smoking, drinking, physical activity, family history of hypertension, BMI, FPG, and LDL-C.
LDL-C, low-density lipoprotein cholesterol; non-HDL-C, non-high-density lipoprotein cholesterol; OR, odds ratio; CI confidence interval; Ref, reference; SD, standard deviation; BMI, body mass index; FPG, fasting plasma glucose.

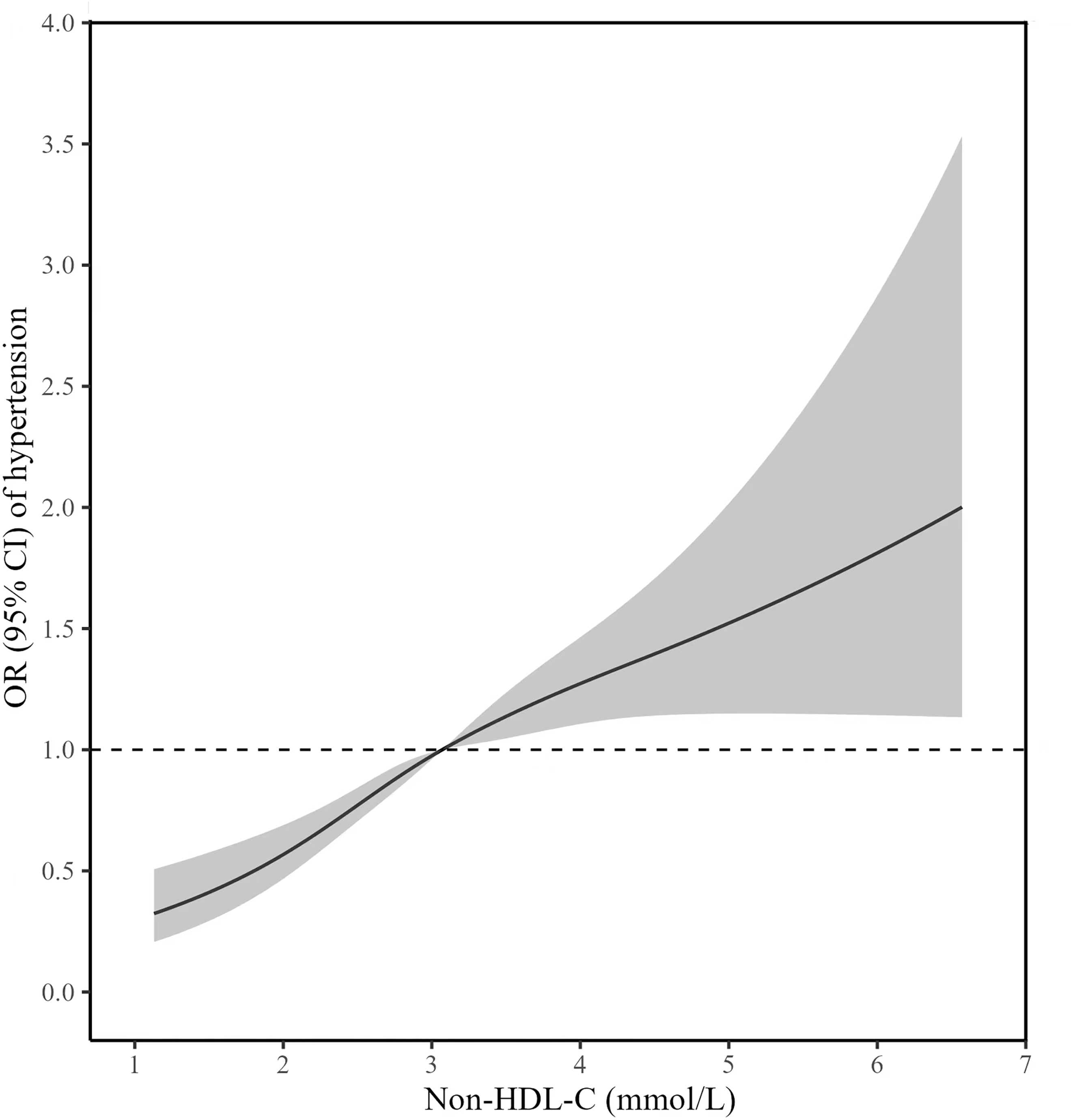

### Subgroup analysis and sensitivity analysis

A significant interaction was identified between non-HDL-C concentration and age for the risk of incident hypertension (*P* _interaction_ < 0.001), with stronger associations between non-HDL-C and risk of incident hypertension in individuals under 60 years versus those over 60. Additionally, a significant interaction was observed between alcohol and non-HDL-C with risk of incident hypertension (*P* _interaction_ < 0.05), suggesting a more obvious association between non-HDL-C and incident hypertension among drinkers versus non-drinkers. Further, a significant interaction was identified between non-HDL-C and a family history of hypertension (*P* _interaction_ < 0.001). Despite the absence of significant interactions between non-HDL-C and other potential hypertension hazard factors in our findings, we found substantial positive associations in males, smokers, and individuals with low to moderate level of physical activity (Figure 2).

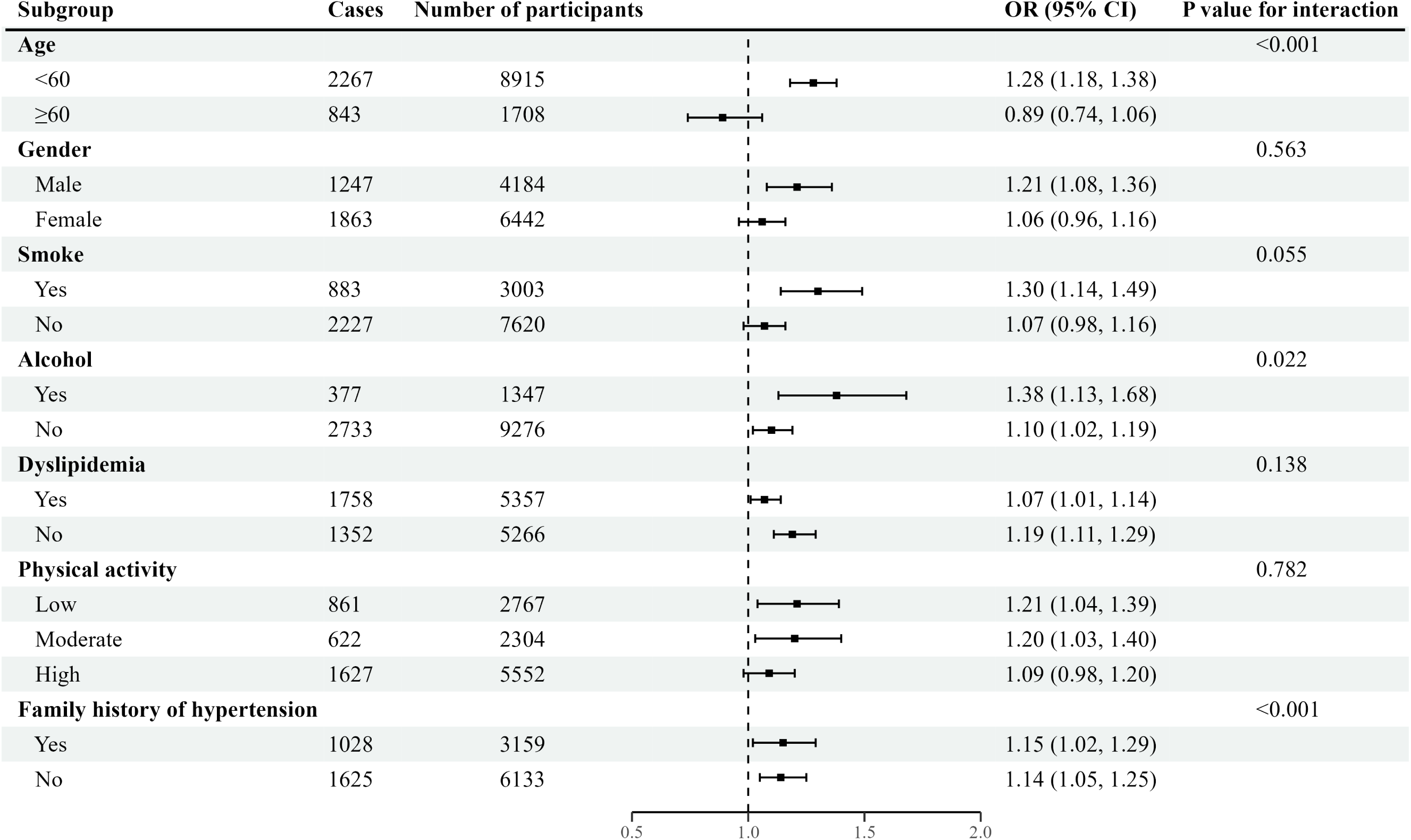

Patients with prehypertension, malignancy, end-stage renal disease, cardiovascular disease, and dyslipidemia at baseline were excluded to enable sensitivity analysis. Sensitivity analysis showed strong overall results, in alignment with the primary results (Supplementary table 2).

### Association between varying levels of LDL-C and non-HDL-C and risk of incident hypertension

The joint association of LDL-C and non-HDL-C for primary outcomes is shown in Table 3. With ideal level of LDL-C (<2.6 mmol/L) and non-HDL-C (<3.4 mmol/L) as the reference group, the risk of incident hypertension increased to an OR of 1.56 (95% CI: 1.23-1.98) for individuals with LDL-C <2.6 mmol/L and non-HDL-C ≥3.4 mmol/L. For those with LDL-C ≥2.6 mmol/L and non-HDL-C <3.4 mmol/L, the risk slightly increased, with an OR of 1.05 (95% CI: 0.91-1.22). When both of them exceeded their respective ideal levels, the risk of incident hypertension increased to an OR of 1.34 (95% CI: 1.20-1.49). Further, there were no discernible multiplicative or additive interactions between LDL-C and non-HDL-C on risk of incident hypertension (multiplicative effect: OR (95% CI): 1.05, 0.79-1.40; additive effect: RERI = 0.06, 95% CI: −0.26, 0.41; Supplementary table 1, model 3).

**Table 3.** Association of different levels of LDL-C and non-HDL-C with risk of incident hypertension.

| LDL-C level<br>(mmol/L) | Non-HDL-C level<br>(mmol/L) | Cases | Number of<br>participants | Cumulative<br>incidence (%) | OR (95% CI) |  |  |
| --- | --- | --- | --- | --- | --- | --- | --- |
|  |  |  |  |  | Model 1 | Model 2 | Model 3 |
| <2.6 | <3.4 | 1404 | 5615 | 25.04 | 1.00 (ref) | 1.00 (ref) | 1.00 (ref) |
|  | ≥3.4 | 139 | 372 | 37.37 | 1.79 (1.44-2.23) | 1.72 (1.38-2.15) | 1.56 (1.23-1.98) |
| ≥2.6 | <3.4 | 393 | 1345 | 29.22 | 1.24 (1.09-1.41) | 1.14 (0.99-1.30) | 1.05 (0.91-1.22) |
|  | ≥3.4 | 1174 | 3292 | 35.66 | 1.66 (1.51-1.83) | 1.46 (1.33-1.61) | 1.34 (1.20-1.49) |

Model 1: unadjusted
Model 2: adjusted for age and gender.
Model 3: model 2 + further adjusted for marital status, education, smoking, drinking, physical activity, family history of hypertension, BMI, and FPG.
LDL-C, low-density lipoprotein cholesterol; non-HDL-C, non-high-density lipoprotein cholesterol; OR, odds ratio; CI confidence interval; Ref, reference; BMI, body mass index; FPG, fasting plasma glucose

## Discussion

To our knowledge, this is the first prospective cohort study to investigate the association of non-HDL-C level with risk of incident hypertension, categorized according to ideal levels of non-HDL-C and LDL-C to assess their distinct influences on risk of incident hypertension. Group 4 was related to a 79% increased risk of incident hypertension compared to group 1, and there was a nonlinear dose-response relationship between serum concentrations of non-HDL-C and risk of incident hypertension, with each additional SD of non-HDL-C corresponding to a 41% heightened risk of incident hypertension. Additionally, compared to ideal levels of LDL and non-HDL-C, those with ideal LDL-C but high blood concentration of non-HDL-C had a 56% greater risk of incident hypertension; however, no correlation was observed when non-HDL was ideal and LDL-C was suboptimal.

In line with current guidelines, the utilization of lipid-lowering drugs should prioritize LDL-C as the primary therapeutic target, with non-HDL-C concentrations as the secondary goal, given that dyslipidemia is a contributing factor to the risk of incident hypertension ^24,25^. Increasing evidence suggest however that non-HDL-C is superior to other standard lipid markers, including LDL-C, and has been demonstrated to be a more valid risk indicator in prevention studies ^26–28^. Non-HDL-C, which can be measured without fasting and has critical values that are simple to calculate, is unaffected by TC level, making it a more reliable indicator than LDL-C when triglycerides are elevated, and it is also easier to explain in the context of health education and risk management ^29–31^. Our results indicate an active association of non-HDL-C with risk of incident hypertension, aligning with earlier studies. Similarly, a cross-sectional study of 117,056 Chinese adults without type 2 diabetes mellitus found a positive association between non-HDL-C and risk of incident hypertension, with a 6.1% increase per SD of non-HDL-C ^14^. Additionally, a prospective study involving 1,019 men showed that males with the highest levels of non-HDL-C had a 36% greater risk of incident hypertension ^5^.

The possible mechanism of serum non-HDL-C leading to hypertension is as follows. First, non-HDL-C’s lipid components possess the ability to reach blood vessels through the wall, resulting in endothelial dysfunction and decreased nitric oxide generation, which in turn decreases vascular dilation capacity and raises vascular tension ^32^. Second, non-HDL-C promotes the release of inflammatory factors through oxidative stress reactions, damaging the vessel wall and causing it to thicken and lose elasticity, thereby raising blood pressure ^33,34^. Third, abnormal blood lipids can reduce the release of substances that dilate blood vessels while increasing the release of vasoconstrictor substances such as endothelin ^35^. Finally, non-HDL-C may cause the renin-angiotensin-aldosterone system to become activated, which would aggravate the development of hypertension by causing water and sodium retention as well as vascular contraction ^36^. These mechanisms interact with each other, driving the pathogenesis of hypertension.

We are investigating additional lipid management tactics beyond just lowering LDL-C as the main treatment, aiming to provide extra benefit when preventing and managing disease risk. The study indicates that non-HDL-C exhibits greater significance than LDL-C for evaluating risk of incident hypertension, as more-than-ideal non-HDL-C concentrations greatly raise the risk of incident hypertension, even if LDL-C level is optimal, whereas the risk does not rise significantly if the non-HDL-C level is ideal. This result is similar to those of earlier studies on outcomes like diabetes, all-cause mortality, and heart-related incidents ^27,37,38^. A notable finding is that the risk of incident hypertension was lower when both of them were non-ideal than when only non-HDL-C was non-ideal. This may be due to the fact that when non-HDL-C is increased and LDL-C is lowered, residual cholesterol (RC) is also raised, with some previous studies showing that RC level is associated with an increased risk of incident hypertension. Multiple previous studies have found that higher RC concentration is linked to an added risk of incident hypertension ^39,40^. On the one hand, RC penetrates the inner wall of the artery and accumulates, stimulating monocytes and macrophages to release pro-inflammatory factors, promoting free fatty acid release and foam cell formation, thereby causing endothelial dysfunction and inflammation ^41,42^. This chronic inflammation may trigger oxidative stress, exacerbating impairment of arterial function ^33^. On the other hand, RC stimulates aldosterone secretion, resulting in sodium conservation as well as elevated blood volume, and thereby raising blood pressure and fostering atherosclerosis ^43,44^.

The study showed a strong association between non-HDL-C and risk of incident hypertension among individuals under 60, males, smokers, drinkers, and those with low to moderate physical activity. Changes with age, lower basal metabolic rate, increased incidence of chronic diseases, and changes in diet may mask or alter the relationship between non-HDL-C level and risk of incident hypertension ^45,46^. In addition, male hormones such as testosterone are affected, which may increase lipid levels and promote the development of atherosclerosis ^47^. Smoking and drinking both damage blood vessel walls, increase blood viscosity, and promote inflammatory responses, while these factors work together with raised concentrations of non-HDL-C to increase the risk of incident hypertension ^48,49^. Simultaneously, levels of low to moderate physical activity may not be sufficient to enhance lipid metabolism or offset the detrimental impacts of such unhealthy lifestyles, resulting in the notable association of non-HDL-C with risk of incident hypertension ^50^.

This is the first prospective cohort study to thoroughly explore the association between blood non-HDL-C concentrations and risk of incident hypertension in rural Chinese adults while also classifying LDL and non-HDL-C into ideal levels, using a large sample and rigorously controlling variables. Still, the study has some limitations. First, although the method for estimating TC is considered accurate, biases in HDL-C measurement could influence the precision of non-HDL-C concentration calculations, especially in the presence of extremely high triglyceride levels. Such discrepancies may introduce bias into the computation of non-HDL-C. Second, the findings probably will not be applicable to other populations since the study was in a rural Chinese setting. Further studies across different populations are needed. Lastly, although we controlled for confounding variables with 4 models, unseen factors like psychological factors could have affected the outcomes.

## Conclusion

This study indicates that higher baseline a non-HDL-C level is nonlinearly and positively associated with risk of incident hypertension among rural Chinese adults. Notably, this correlation was independent of LDL-C level, suggesting that further reductions in non-HDL-C, based on already controlled LDL-C, were associated with a further decrease in the risk of incident hypertension. To validate our findings, further studies with larger populations of varying ethnic populations are required.

## Data Availability

Contact the corresponding author for all data relating to this study upon reasonable request.

## Non-standard Abbreviations and Acronym

AHA: American Heart Association
AP: proportion attributable to interaction
BMI: body mass index
CVDs: cardiovascular diseases
FPG: fasting plasma glucose
LDL-C: low-density lipoprotein
Non-HDL-C: non-high-density lipoprotein cholesterol
RCS: restricted cubic splines
RC: residual cholesterol
RERI: relative excess risk due to interaction
SI: synergy index
TC: total cholesterol
TG: triglycerides

## Acknowledgement

We acknowledge all the authors, particularly Prof. Dongsheng Hu for methodological advice and help in modifying the manuscript.

## Supplemental Material

Tables S1-S2

Figures S1

